# Markers of brain and endothelial Injury and inflammation are acutely and sex specifically regulated in SARS-CoV-2 infection

**DOI:** 10.1101/2021.05.25.21257353

**Authors:** Jude Savarraj, Eun S. Park, Gabriela Copo, Sarah Hinds, Diego Morales, Hilda Ahnstedt, Atzhiry Paz, Andres Assing, Shivanki Juneja, Eunhee Kim, Sung-min Cho, Aaron Gusdon, Pramod Dash, Louise McCullough, H Alex Choi

## Abstract

**Objective:** To investigate brain injury markers (BIM), endothelial injury markers (EIM) and cytokine/chemokine (CC) markers of systemic inflammation in coronavirus disease 2019 (COVID-19) and across sex.

**Methods:** Plasma samples from 57 subjects at <48 hours of COVID-19 hospitalization, 14 subjects at 3 months of COVID-19 hospitalization and 20 matched controls were interrogated for the levels of six BIMs - including GFAP, S100B, Syndecan-1, UCHLI, MAP2 and NSE, two EIMs – including sICAM1 and sVCAM1 and thirty-eight CCs. Statistical and bioinformatics methods were used to measure differences in the marker profiles across (a) COVID-19 vs controls and (b) men vs women.

**Results:** Three BIMs: MAP2, NSE and S100B, two EIMs: sICAM1 and sVCAM1 and seven CCs: GRO IL10, sCD40L, IP10, IL1Ra, MCP1 and TNFα were significantly (p<0.05) elevated in the COVID-19 cohort compared to controls. Two CCs: MDC and MIP1α were significantly lower in the COVID-19 cohort. Bioinformatics analysis reveal a stronger positive association between BIM/CC/EIMs in the COVID-19 cohort. Analysis across sex revealed that several BIMs and CCs including NSE, IL10, IL15 and IL8 were significantly (p<0.05) higher in men compared to women. Men also expressed a more robust BIM/ EIM/CC association profile compared to women. At 3 months, BIMs and CCs were not significantly different in the COVID-19 cohort compared to controls.

**Conclusion:** The acute elevation of BIMs, CCs, and EIMs and the robust associations among them at COVID-19 hospitalization suggest that brain injury is mediated by endotheliopathy and inflammation. Higher BIM and inflammatory markers in men additionally suggest that men are more susceptible to the risk compared to women.

## Introduction

Neurologic complications after COVID-19 have been reported in both the acute and chronic period^1,2^. Neurologic manifestations including ischemic and hemorrhagic stroke (which are direct acute medical complications of COVID-19) have been reported. But neurologic manifestations without clear etiology including encephalopathy, pain, chronic fatigue and cognitive dysfunction have also been reported demonstrating the wide impact of COVID-19 on the nervous system^3–8^. Human coronaviruses (HCoV) are neuroinvasive and neurotropic by nature and may contribute to both short-and long-term neurological disorders^9–11^. SARS-CoV (of the SARS epidemic in early 2000s) has been shown to affect the CNS^12^ and viral strains have been identified in post-mortem human brain tissue^12,13^. Our understanding of the effect of COVID-19 on brain injury is only beginning to be developed. COVID-19 triggers multiple pathological mechanisms -including an acute inflammatory response and endotheliopathy -and the relationship of these mechanisms with brain injury is unclear. Further, it is not known whether the BIMs continue to be elevated in the long-term after COVID-19.

Sex differences in COVID-19 are prominent, with men having higher hospitalization rates and mortality rates-an effect seen globally^14,15^. Sex differences in the immune responses have been shown to underlie the increase in mortality seen in men during the acute disease course^16,17^. It is uncertain whether the increase in peripheral inflammation seen in accompanied by an increase in brain injury markers as well.

We investigated brain injury in COVID-19 patients by measuring brain injury markers (BIMs) in the acute (<48h) and chronic phase (3 months) after hospitalization. Several BIMs have been established in neurologic diseases and have been reported as surrogate markers of neuronal and astrocytic injury in non-neurological diseases like HIV^18–20^, cardiac arrest^21–23^ and sepsis^24^ as well. Among them, we measured six validated BIMs including glial fibrillary acidic protein (GFAP), neuron specific enolase (NSE), S100B, ubiquitin carboxyl-terminal hydrolase isozyme L1 (UCHL1), Syndecan-1 and microtubule-associated protein 2 (MAP 2) in plasma drawn from hospitalized COVID-19 subjects and matched controls. In addition to BIMs, we measured two endothelial injury markers (EIMs) (including *Intercellular Adhesion Molecule 1* (ICAM-1) and *Vascular Cell Adhesion Molecule 1* (VCAM-1)) and several cytokine/chemokine markers (CCs) of systemic inflammation to investigate if endothelial injury and inflammation are associated with BIMs. The levels of the markers were measured by ELISA and multiplex assays, and the association between COVID-19 and controls were characterized using statistical and bioinformatics approaches. The differences in the levels of markers between men and women were investigated.

## Methods

### Study population and patient inclusion and exclusion criteria

This is a prospective study of COVID-19 patients admitted and hospitalized at the Memorial Herman Hospital, Houston, Texas, USA. Inclusion criteria were laboratory-confirmed SARS-CoV-2 infection by real-time polymerase chain reaction, written informed consent from the patient or surrogate and age ≥ 18 years of age. Exclusion criteria were inability to complete long-term follow-up, severe functional disabilities before hospital admission for COVID-19 (defined by pre-admission modified Rankin Score(mRS)^25^ >1), history of pulmonary complications (including resection and transplant), pre-existing systemic diseases which would impact long term outcomes (including stroke, myocardial infarction, pulmonary disease requiring home oxygen, chronic renal failure necessitating hemodialysis and malignancy), documented neurologic and psychiatric disorders, prisoners and pregnant women. Patients were categorized as mild (nasal cannula with <5 liters of O_2_ and <5 days of hospitalization), moderate (nasal cannula with >5 liters of O_2_ or heat high flow cannula and > 5 days of hospitalization) and severe (on ventilator or expired). Plasma samples from 24 non-neurological subjects who were enrolled at the *UT Physician Cardiology* clinic were used as controls. The controls were matched to the COVID-19 cohort for age, sex and co-morbidities (including diabetes and hypertension). This study was approved by the ‘Institutional Review Board’ (IRB No: HSC –MH-17-0452) at The University of Texas Health Science Center at Houston, Houston, Texas.

### Samples

Blood was drawn by venipuncture, collected into sterile vacutainers, and immediately placed on ice. For processing of plasma, the tubes were centrifuged at 1200 x *g* for 10 minutes at 4°C followed by a second centrifugation at 10,000 x *g* for 10 minutes at 4°C to generate platelet poor plasma. Plasma were then aliquoted and stored at -80°C until the samples were analyzed.

### ELISA and multiplex analysis

BIMs were measured using the following ELISA kits: GFAP (Cat No: NS830, Sigma-Aldrich), NSE (Cat No: 420-85, IBL America), S100B (Cat No: EZHS100B-33K, MilliporeSigma), UCH-L1 (EH475RB, ThermoFisher), syndecan-1(RAB0736-1KT, MilliporeSigma) and MAP2 (EKU05950, BIOMATIK). The EIMs (including ICAM-1 and VCAM-1) were measured using a premixed multiplex assay (Cat. #HCVD2MAG-67K, MilliporeSigma). Systemic levels of CCs were measured using a 38-plex premixed immunological multiplex assay (HCYTMAG-60K-PX38, MilliporeSigma, Billerica, MA). CCs included CD40L, EGF, Eotaxin/CCL11, FGF-2, Flt-3 ligand, Fractalkine, G-CSF, GM-CSF, GRO, IFN-α2, IFN-γ, IL-1α, IL-1β, IL-1ra, IL-2, IL-3, IL-4, IL-5, IL-6, IL-7, IL-8, IL-9, IL-10, IL-12 (p40), IL-12 (p70), IL-13, IL-15, IL-17A, IP-10, MCP-1, MCP-3, MDC (CCL22), MIP-1α, MIP-1β, TGF-α, TNF-α, TNF-β And VEGF. All experiments were performed according to manufacturer’s protocol. The levels of all markers are reported as pg/ml.

### Statistical analysis

Descriptive statistics were calculated for demographic variables and protein levels in control and COVID-19 subjects. To describe differences in demographics, χ2-test, Fisher’s exact test, student’s t-test, and the Mann-Whitney U test were used where appropriate. The Mann-Whitney U test was used to test for differences in protein levels across different groups A *p*-value of ≤□0.05 was considered statistically significant (two-tailed). The levels of the markers are reported as mean ± standard error (SE). All statistical analyses were performed using open-source software packages in R (v3.1.3)

### Bioinformatics and Chord diagrams

To elucidate the relationship between the BIMs, EIMs and CCs, we employed chord diagrams. Measurements of markers were normalized via Box-Cox transformations and Pearson’s correlation coefficient (PCC) was calculated (R v3.1.3) for each pair of the measurements. Only significant (p<0.05) associations were retained were used to construct the dendrograms and the chord diagrams. The R package *circlize*^26^ was used to construct the chord diagrams.

### Modified Rankin assessment

The functional status of the patient was quantified using the modified Rankin’s scale (mRS) which is a 0-6 point grading scale of the patient’s functional status^27^. ‘0’: no deficits, ‘1’: presence of symptoms but no significant disability, ‘2’: a slight disability but independent, ‘3’: moderate disability requiring some help but able to walk without assistance, ‘4’: a moderately severe disability and unable to walk and attend to bodily needs without assistance, ‘5’: a severe disability where the patient requires constant nursing care and attention and, ‘6’: being dead. For the purposes of this study, we dichotomized patients into two groups; good (mRS≤3) and poor (mRS≥4) outcomes. The mRS at discharge was assessed by the attending neurointensivist taking care of the patient.

## Results

### Patient characteristics

Samples from 57 subjects and 20 controls subjects were included in the study. Demographics and clinical outcomes of the COVID-19 subjects and controls are presented (**Table 1**). There were no significant differences in age, sex and race distribution between the two groups. The COVID-19 cohort had a significantly more Hispanic subjects compared to the control (56% vs 10%, *p<0*.*01*). There were no differences in the history of hypertension, diabetes and coronary heart disease between the two groups. However, the COVID-19 group had more obese subjects compared to controls (49% vs 20%, *p=0*.*03*).

**Table 1:** Demographics and past medical history.

| Variables | Control<br>N=20 | COVID-19<br>N=57 | *p* | COVID-19 at<br>3 months<br>N=14 | *p* |
| --- | --- | --- | --- | --- | --- |
| **Demographics** |  |  |  |  |  |
| Age (mean,sd) | 61(15) | 53(18) | 0.07 | 51(14) | 0.07 |
| Sex, female (n, %) | 11(55) | 24(42) | 0.4 | 6(40) | 1 |
| Race (n, %) |  |  |  |  |  |
| White | 16(80) | 35(61) | 0.17 | 11(78) | 0.7 |
| African-American | 3(15) | 15(26) | 0.37 | 2(14) | 1 |
| Others | 1(5) | 7(12) | 0.67 | 1(7) | 1 |
| Ethnicity, Hispanic (n, %) | 2(10) | 32(56) | **<0.01** | 12(85) | **<0.01** |
| **Past Medical History** |  |  |  |  |  |
| Hypertension (n, %) | 5(25) | 24(42) | 0.28 | 5(35) | 0.7 |
| Diabetes (n, %) | 4(20) | 24(42) | 0.16 | 5(35) | 1 |
| CAD (n, %) | 5(25) | 7(12) | 0.16 | 0(0) | 0.06 |
| Obesity (n, %) | 4(20) | 28(49) | **0.03** | 4(28) | 0.41 |
| SD: Standard Deviation; CAD: Coronary artery disease |  |  |  |  |  |

### BIMs after COVID-19 hospitalization

At <48 hours of COVID-19 hospitalization, the mean levels of MAP2 (68 ± 7.5 vs 26 ± 3.9, pg/ml, p<0.01), NSE (16.9 ± 3.2 vs 4.9 ± 0.59, pg/ml, *p*<0.01) and S100B (113 ± 16 vs 43 ± 7, pg/ml, *p*<0.01) were significantly higher compared to controls (**Figure 1A**). In the COVID-19 cohort, the average levels of MAP2, NSE and S100B were 160%, 245% and 162% higher than controls. However, at 3 months after hospitalization, the levels of these markers were not significantly different from controls (**Figure 1A**).

**Figure 1:**
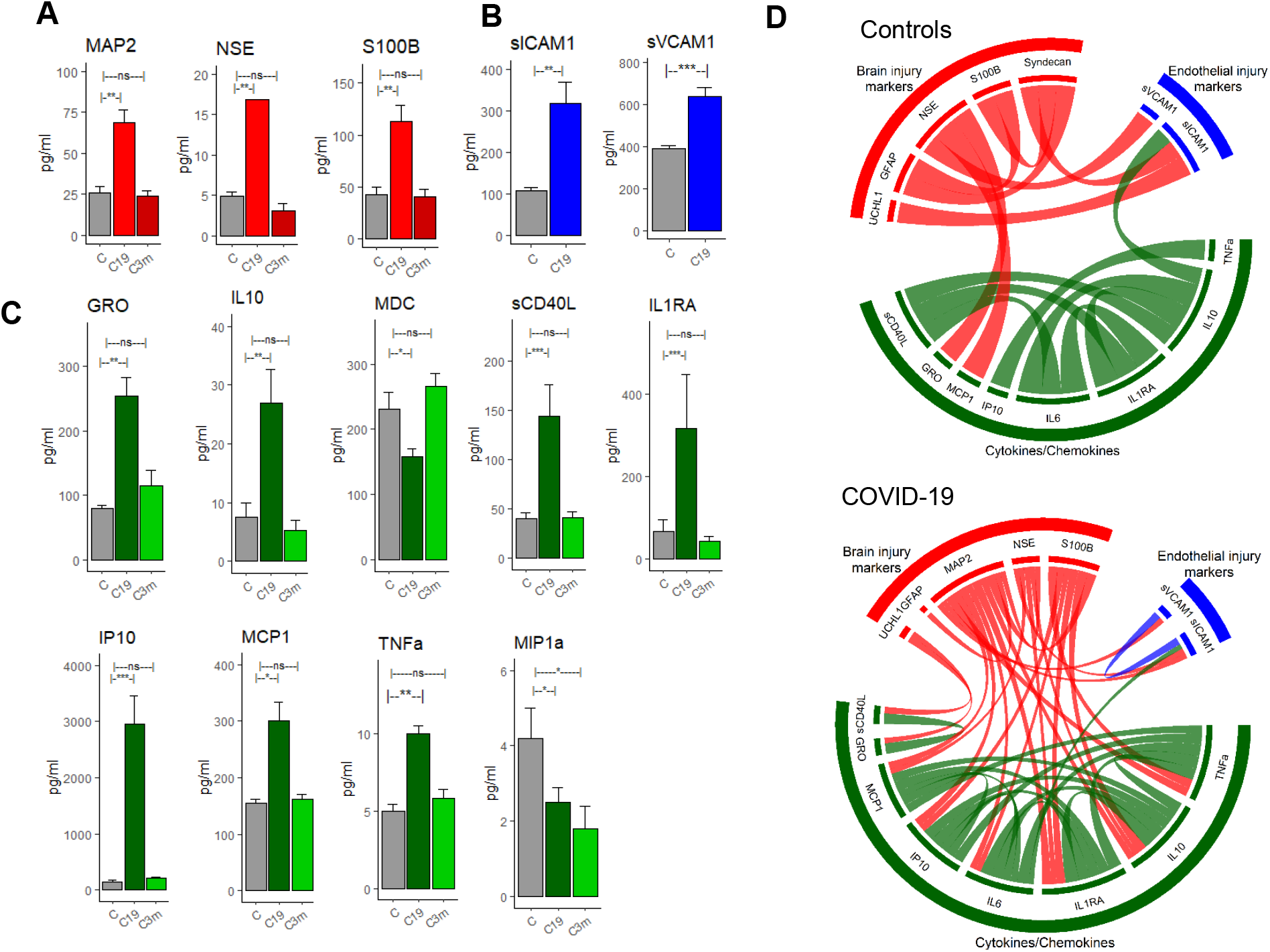
Difference in BIM, EIM and CC profile across COVID-19 subjects and matched controls. (A) BIMs including MAP2, NSE and S100B were significantly (p<0.05) higher immediately after COVID-19 hospitalization and returned to baseline values at 3 months (B) sICAM1 and SVCAM1 levels were significantly higher after COVID-19 hospitalization (C) Several inflammatory markers including GRO, IL10, sCD40L, IL1RA, IP10, MCP1 and TNFa were significantly higher after COVID-19. MDC and MIP1a were significantly lower after COVID-19. Except MIP1a, all CC markers returned to baseline levels at 3 months. MIP1a continue to remain lower at 3 months (D) Chord diagram reveals significant (p<0.05, Fishers’ test) differences in interactions between the BIM, EM, and CCs across COVID-19 and control groups.

### EIMs and CCs after COVID-19 hospitalization

At the acute phase (<48h), the levels of the EIMs and most CCs were significantly higher in the COVID-19 cohort compared to controls. EIMs including sICAM1 (317 ± 51 vs 108 ± 7.5, pg/ml, p<0.001) and sVCAM1 (637 ± 45 vs 390 ± 16, pg/ml, p<0.01) were significantly higher in COVID-19 compared to controls (**Figure 1B**). Of the 38 CCs investigated, 7 CCs were significantly higher and 2 CCs were significantly lower in COVID-19 (**Figure 1C**). The 7 CCs that were significantly higher were GRO (254 ± 29 vs 79 ± 5.19, pg/ml, p<0.01), IL10 (27 ± 5.7 vs 7.4 ± 2.5, pg/ml, p<0.01), sCD40L (144 ± 32 vs 40 ± 6, pg/ml, p<0.0001), IL1Ra(318 ± 131 vs 68 ± 28, pg/ml, p<0.01), IP10 (2952 ± 512 vs 148 ± 20, pg/ml, p<0.001), MCP1 (300 ± 34 vs 154 ± 8, pg/ml, p< 0.05) and TNFa (10 ± 0.5 vs 5± 0.4, pg/ml, p<0.01). The two CCs that were significantly lower in the COVID-19 cohort were MDC (158 ± 12 vs 231 ± 26, pg/ml, p<0.05) and MIP1a (2.5 ±0.4 vs 4.2 ± 0.8, pg/ml, p<0.05). At 3 months after COVID-19 hospitalization, the levels of BIMs and CCs (with the exception of MIP1a) were similar to the levels of the control cohort. We did not measure EIM levels at 3 months.

### The association with inflammatory cytokines and COVID-19

Bioinformatics (Chord diagrams) analysis reveal distinct association profile between the COVID-19 and control cohort (**Figure 1D**). Within the COVID-19 cohort, we observed high positive correlations between the BIM and CCs indicative of a brain injury mechanism triggered by systemic inflammation. While IL6 levels were not significantly higher in the COVID-19 cohort, as previous studies have found they trended higher in men. In our cohort, we observed IL6 to highly correlated with other CCs and BIMs (**Figure 1D**). Several CCs and notably MCP1, IP10, IL6, IL1Ra, IL10 and TNFa had more associations (**Figure 1D, COVID-19**) with the BEM markers suggesting that these CCs are the inflammatory markers driving brain injury.

### Severity and outcomes

We examined the levels of BIMs, EIMs and CCs across COVID-19 severity (mild, moderate and severe). The levels of BIM were not significantly different across severity. However, one EIM (sICAM1) and two CCs (SCD40L and IL1RA) were significantly higher in subjects with high severity (**Figure 2A**). None of the BIMs or EIMs were predictive of functional outcomes at discharge. Two CCs, Eotaxin and MIP1b were significantly higher (p<0.05) in subjects who proceeded to have poor outcomes (**Figure 2B**). When adjusted for age and sex, only MIP1b was found to be independently associated with functional outcomes. MDC was significantly (p<0.05) lower in those who proceeded to have poor outcomes. However, when controlled for age and sex, it was not independently associated with outcomes (**Figure 2B**).

**Figure 2:**
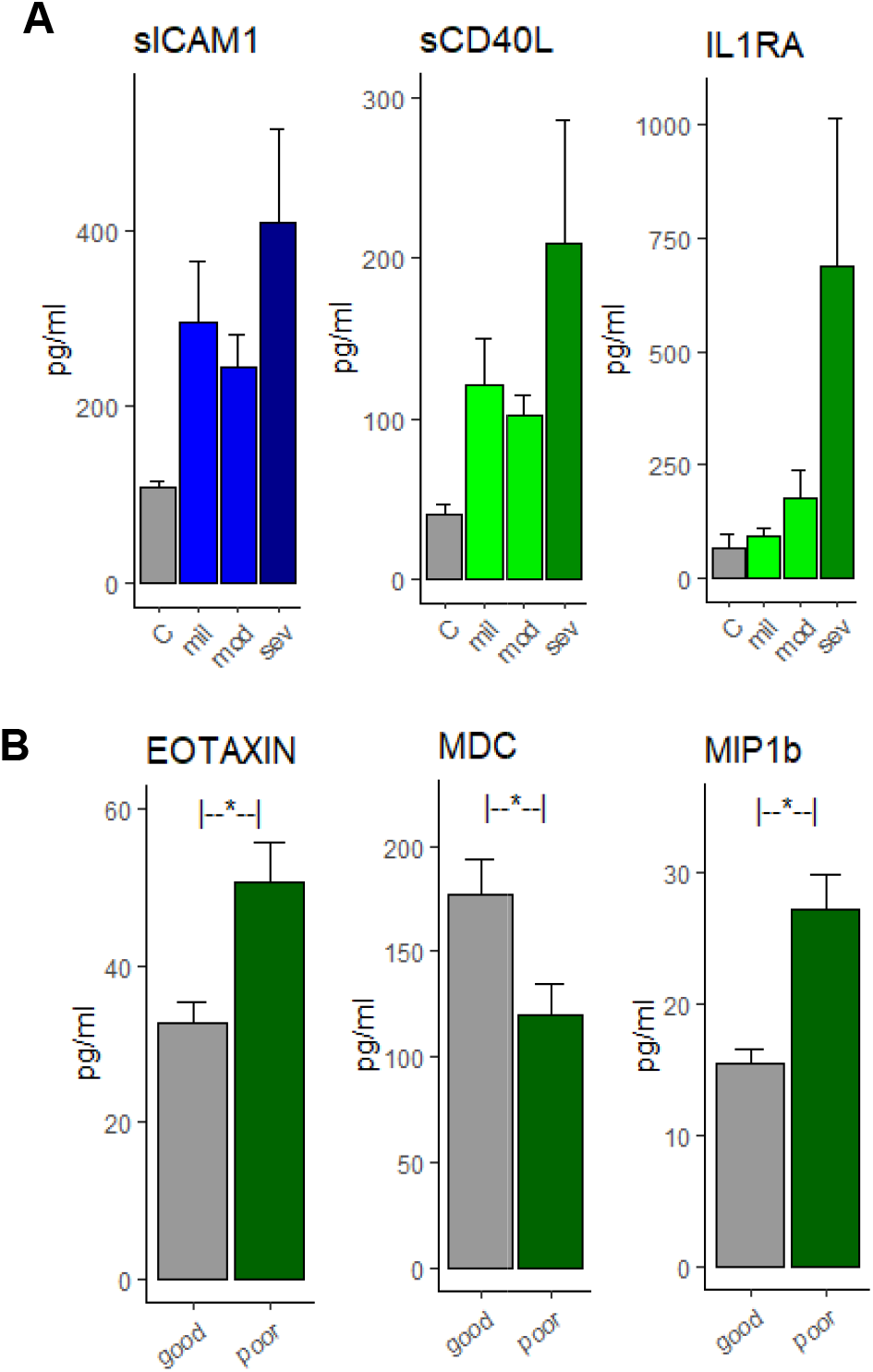
Differences across severity and clinical outcomes. (A) Difference in SICAM1, sCD40L and IL1RA across controls, mild, moderate and severe COVID groups. (B) Difference in CC levels across functional outcomes at discharge.

### Sex Differences

Among the BIMs, only the levels of NSE was significantly higher in men compared to women (7.95 ± 0.9 vs 22.9 ± 5.8, *p*=0.01). BIMs including S100B, tau and MAP2 were higher in men; however, these differences were not statistically significant (**Figure 3A**). The levels of the EIMs were not significantly different across men and women (data not shown). Four CCs were significantly higher in men compared to women: IL10 (35 ± 10.2 vs 14.4 ± 2.3, p=0.05), IL15 (5 ± 1.1 vs 2.4 ± 0.6, p=0.4), IL8 (32 ± 9.2 vs 11.4 ± 3.06, p=0.04) and MIP1a (3.3 ± 0.7 vs 1.4 ± 0.4, p=0.4). Four CCs including MIP1b, FGP2, GMCSF and IL6 were near significantly (p<0.1) higher in men compared to women (**Figure 3B**). The BIM-EIM-CC interaction profile reveal stark differences between men than women (**Figure 3C**). Men had a higher number of correlations between the BIM-EIM-CC and the correlation profile was significantly different compared to women (p<0.05, Fisher’s test). There were more interactions between the CCs and BIMs suggestive of a higher degree of inflammation induced brain injury in men. Additionally, in women, the EIMs (sICAM1 and SVCAM1) were not significantly correlated with the CCs or BIMs. However, in men, the EIMS were highly correlated with CCs and BIMs (**Figure 3C**).

**Figure 3:**
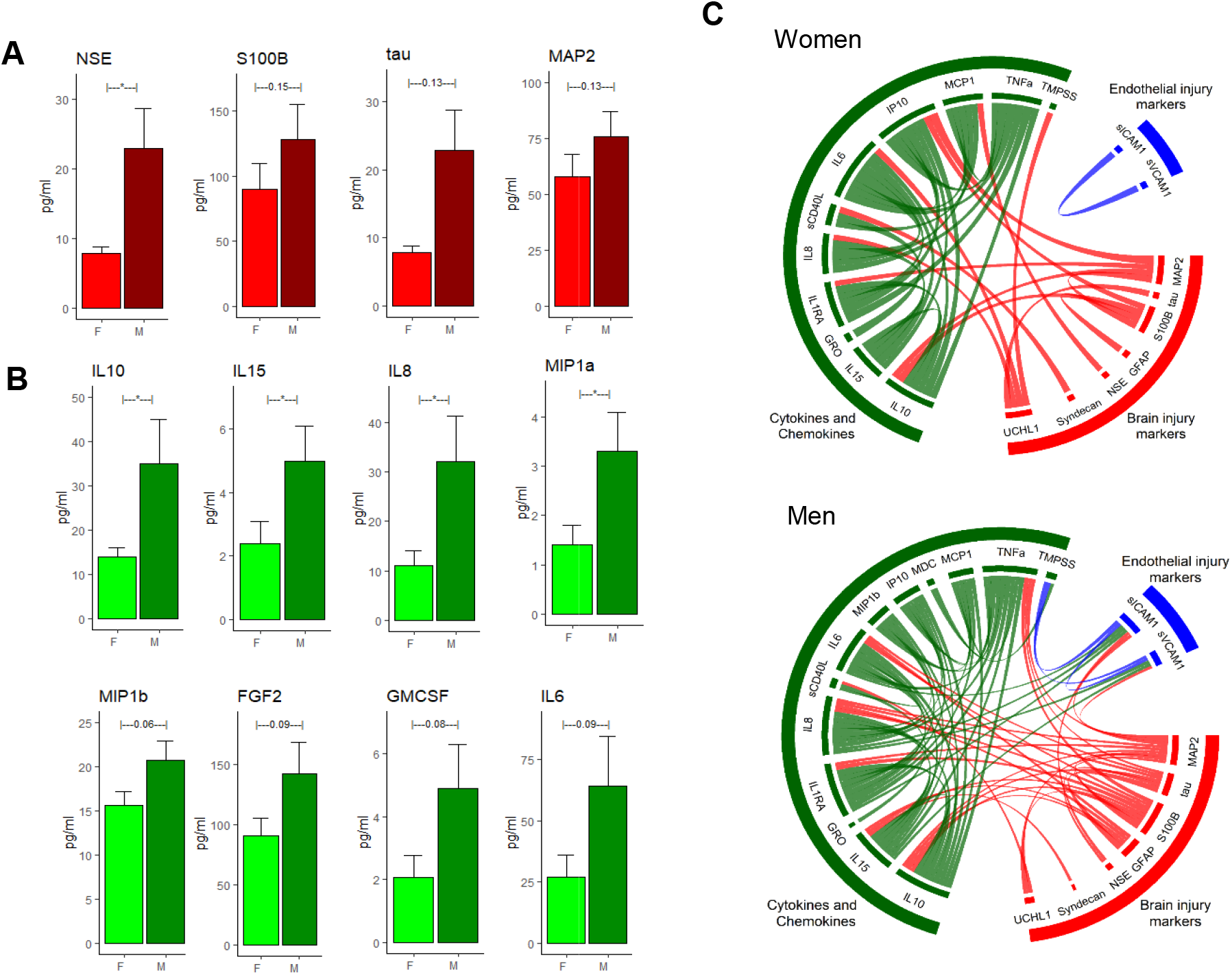
Sex difference in BIM and CC profile across COVID-19 subjects. (A) NSE was significantly (p<0.05) higher after COVID-19. S100b, tau and MAP2 were higher but differences was not statistically significant (B) IL10, IL15, IL8 and MIP1a were significantly (p<0.05) in males compared to females. MIP1b, FGF2, GMCSF and IL6 indicated a non-significant trend of being elevated in males (C) The BIM-EM-CC interactions reveal that males have a much robust interactions compared to females. There were no strong correlations between the EIMs and BIM/CCs in females. There were strong associations between EIM, CC and BIM in males.

## Discussion

We investigated the profile of brain injury markers, endothelial injury and inflammatory markers after COVID-19 disease. We found that levels of BIMs, specifically MAP2, NSE and S100β were significantly elevated in the acute phase of COVID-19, with levels returning to normal at 3 months after hospitalization. We found that two EIMs, sICAM1 and sCAM-1, were elevated after COVID-19 infection with robust interactions with BIMs and IL6. Additionally, we identified a host of inflammatory markers that are elevated after acute COVID-19 infection and most return to normal at 3 months. Interestingly we found BIM in men were significantly higher than in women suggesting a strong sex dependent biological mechanism underlying the interactions between systemic inflammation, endothelial damage and brain injury after COVID-19.

The neuro-invasive and neurotropic nature of human coronaviruses (HCoV) are known, and they may contribute to both short-and long-term neurological disorders ^9–11^. The neuroinvasion of HCoVs confirmed by the presence of viral RNA in the human CNS adds credence to this hypothesis^28^. Acute neurologic and psychiatric complications have been reported after COVID-19 hospitalizations^2^. Neurologic manifestations have been reported in both the central nervous system and the peripheral nervous system. A wide range of neurological manifestations including ischemic stroke, intracerebral hemorrhage, hallucinations, encephalopathy, anosmia and ageusia have been described after SARS-CoV-2 infection^3–8^. Hyperemic and edematous brain tissue and neuronal death have been reported in autopsy cases after COVID-19^29^.

To investigate the effects of COVID-19 on BIMs, we examined systemic levels of six previously validated BIMs – MAP2, NSE, GFAP, S100B, Syndecan-1 and UCHL1. We found that MAP2, NSE and S100B were higher after COVID-19 indicative of brain injury after COVID-19. MAP-2 is a dendritic injury marker and was found to be elevated in both the acute and chronic stages of brain injury^30^. It is a protein that is typically localized in the dendritic regions and is involved in the promotion of microtubule synthesis and cross-linking with other compartments of the cytoskeleton. It is elevated after a traumatic brain injury and was found to be associated with long-term outcomes^30^. NSE is an acidic protease present in neurons and neuroendocrine cells and is an useful indicator of neuronal damage^31^. S100B is a calcium-binding protein that is localized in astrocytic cytoplasm. It has been shown to be elevated systemically especially in traumatic brain injuries^32^ and strokes^33^. Systemic levels of S100B is higher after traumatic brain injury and have been associated with poor long-term outcomes^34–36^. Increased levels of MAP2, NSE and S100B indicates neuronal and astrocytic injury after COVID-19 (**Figure 1A**). Additionally, we found that BIMs (NSE, S100β, tau, MAP2) were higher in men compared to women with COVID-19. It is not clear whether the elevations in BIMs overall and specifically in men have clinical relevance. We deliberately excluded samples from subjects with clear neurologic injury (i.e., traumatic brain injury, ischemic stroke, or hemorrhagic stroke) associated with COVID-19 disease. The elevated BIMs suggest that there is CNS involvement in acute COVID-19 and the clinical ramifications need to be investigated urgently.

Several pro-inflammatory cytokines including IL10, MCP1, TNFa, MDC, GRO, sCD40L, IL1Ra and IP10 were higher after COVID-19 compared to controls confirming the findings of several studies that have reported an increased inflammatory response in COVID-19^37^. We also observed that EIMs (sICAM-1 and sVCAM-1) were higher after COVID-19 infection (**Figure 1B**). Endothelial cells express the ACE-2 receptor – the primary receptor and main route in intracellular entry for SARS-CoV-2^38^.The blood-brain barrier (BBB) via the endothelial tight junctions is critical in maintaining cerebral homeostasis and the breakdown of BBB is implicated in variety of neurological diseases^39^. It has been shown that the BBB is compromised after COVID-19 and that the SARS-CoV-2 spike protein can cross the BBB into brain tissue^40,41^. The endothelial dysfunction might be a direct result of the SARS-CoV-2 infection or due to a reaction to systemic inflammation triggered by the viral infection, or a combination of these factors. ICAM-1 is expressed in the vascular endothelium and is typically induced by IL-1b and TNFα^42^. It has been shown to promote leukocyte adhesion^43^ and effects barrier function and vascular leak^44^. sICAM-1 is the circulating form of ICAM-1 and has been identified as a candidate marker of vascular inflammation^45^. Increased levels of sICAM-1 have been observed in cardiovascular disease including myocarditis^46^ and inflammatory cardiomyopathy^47^. In addition to increased levels of sICAM-1, we found that TNFα was also elevated after COVID-19. In our data, the simultaneous elevation of the pro-inflammatory TNFα and sICAM-1 is suggestive of an inflammatory mediated vascular injury rather than a virally triggered vascular injury (**Figure 1**). In fact, a recent NIH funded post-mortem study examining the brain tissue of deceased COVID-19 subjects reported extensive inflammation and microvascular blood vessel damage, but it is not clear how SARS-CoV-2 is involved in the brain injury and neurological outcome^48^. The findings of this study suggest that the microvascular blood vessel damage is likely caused by the inflammatory response due to the infection, rather than the virus itself.

To elucidate the relationships between the BIMs, EIMs and CCs,0 we employed a bioinformatics visualization tool called the chord diagram and overlaid the significant correlations. In the COVID-19 cohort, the CCs IL6, IL1Ra, IL10, TNFa, MCP1 and IP10 had high associations with MAP2, NSE and S100B. Among the BIMs, MAP2 and S100B had the most associations with the CCs (**Figure 1D**). The relatively higher correlations between the BIMs and CCs in the COVID-19 cohort compared to controls suggest inflammation induced brain injury in COVID-19. It is known that inflammatory diseases are associated with neuronal injury (as in the case of encephalitis after sepsis^49^). Therefore, it is plausible that the extensive cytokine activation by COVID-19 infection is the primary inflammation driver and mediator of neuronal injury.

Interestingly, the levels of BIMs were not different across clinical severity or clinical outcomes. However, sICAM1 and two CCs (sCD40L and IL1Ra) were higher in severe grade subjects (**Figure 2A**) and three CCs (Eotaxin, MDC and MIP1b) were different across clinical outcomes (**Figure 2B)**. After adjusting for age and sex, only MIP1b was independently associated with clinical outcomes. The lack of differences in BIMs levels across severity or clinical outcomes could be a reflection of the time-point at which the samples were drawn rather than a pathophysiological mechanism of the disease process itself as we only analyzed the first sample that was available during hospitalization (obtained at <48 hours after admission) a time-period where typically the heterogeneity in clinical status is low.

The long-term neurological effects of COVID-19 are being increasingly recognized^50^. As more COVID-19 patients are recovering from hospitalization, it is expected that many discharged patients are afflicted with several secondary complications and symptoms^51^. These patients are referred to as “long-haulers” and they are reporting a host of medical symptoms that include functional, cognitive and psychiatric deficits^52–54^. We investigated whether brain injury and inflammation persists at 3-months after hospitalization and found that the systemic levels of BIMs (including MAP2, NSE and S100B) and most inflammatory CCs (except MIPI1a) were similar to the levels observed in controls indicating that there is no ongoing brain injury or higher inflammation at 3 months after COVID-19 hospitalization. It is likely that the neurological sequelae observed in the survivors are due to the consequence of the initial inflammatory cascade at the time of hospitalization rather than an ongoing process at 3 months.

COVID-19 disproportionately affects men more severely compared to women. Preliminary studies have suggested that while the prevalence of infection is the same in men and women, men are more likely to be hospitalized and have a severe course of disease and higher mortality than women^15^. We investigated differences in BIMs and CCs across men and women and we found that NSE was significantly elevated in men compared to women. Additionally MAP2, tau and S100B showed trends towards elevated levels in men (**Figure 3A**). Several CCs including IL15, IL8, MIP1b and IL10 were elevated in men compared to women (**Figure 3B**) suggesting an increased inflammatory response in men. Men had a higher degree of inflammatory interactions (**Figure 3C**) compared to women. Additionally, the interactions between the inflammatory markers (CCs) and the BIMs (red lines) were much dense in men indicating a higher degree of inflammation mediated brain injury in men during the acute stages of COVID-19 (**Figure 3C**). Interestingly, the interactions between the EIMs and the BIMs/CCs were also higher in men compared to women (**Figure 3C**). This suggests that men likely experience a higher degree of inflammation triggered endotheliopathy – an important factor to be considered when developing treatment plans. A number of reasons have been postulated for the differential response across sex to COVID-19. Some studies have shown that men have a higher level of ACE2 receptors, used for SARS-CoV-2 virus entry^55^. Others have postulated that the differences are due to differences in lifestyle factors and co-morbidities between men and women^55,56^. Given our small sample size, we are not powered to determine if these differences in BIMs translate into worse neurologic outcomes. Future studies investigating the differences in the sex profile across the sexes are required.

### Limitations

Our study has a few limitations. First, we included samples only the first available time-point after hospitalization. Typically, the measurements from the first-available time point is a reflection of the patient’s status during the earlier course of disease progress. However, this time-point is also the most accurate as in our hospital; all COVID-19 patients were administered dexamethasone, which could have a confounding effect on the measurements. Second, we have not related the observed BIMs with any long-term clinical outcomes. Third, we are underpowered to detect the effect of any co-morbidities on the neuronal injury.

## Conclusion

Our findings suggest that immediately after COVID-19 hospitalization, BIMs were higher than matched controls. The levels of EIMs and CCs were also higher. The CCs were highly correlated with BIMs and EIMs suggesting inflammatory-driven endothelial dysfunction and brain injury in the acute phase of COVID-19. Further studies are required to clarify the exact mechanisms how COVID-19 induces inflammation and brain injury, sex-specific differences and whether these early increase in injury markers are associated with long-term neurologic outcomes.

## Data Availability

Upon request.

## Acknowledgements

The authors thank all the health care workers and the research personnel who tirelessly cared for the worked were involved in the care and the follow-up of the patients. This work was partly supported by an award from the Huffington Foundation to Dr. Louise D. McCullough. We thank The Biorepository of Neurological Diseases team for their efforts in collecting the samples.

